# Detection and quantification of enteric pathogens in aerosols near open wastewater canals in cities with poor sanitation

**DOI:** 10.1101/2021.02.14.21251650

**Authors:** Olivia Ginn, Lucas Rocha-Melogno, Aaron Bivins, Sarah Lowry, Maria Cardelino, Dennis Nichols, Sachchida Nand Tripathi, Freddy Soria, Marcos Andrade, Mike Bergin, Marc A. Deshusses, Joe Brown

## Abstract

Urban sanitation infrastructure is inadequate in many low-income countries, leading to the presence of highly concentrated, uncontained fecal waste streams in densely populated areas. Combined with mechanisms of aerosolization, airborne transport of enteric microbes and their genetic material is possible in such settings but remains poorly characterized. We detected and quantified enteric pathogen-associated gene targets in aerosol samples near open wastewater canals (OWCs) or wastewater-impacted surface waters and control sites in La Paz, Bolivia; Kanpur, India; and Atlanta, USA via multiplex reverse-transcription qPCR (37 targets) and ddPCR (13 targets). We detected a wide range of enteric targets, some not previously reported in extramural urban aerosols, with more frequent detections of all enteric targets at higher densities in La Paz and Kanpur near OWCs. We report density estimates ranging up to 4.7 × 10^2^ gc per m^3^_air_ across all targets including heat stabile enterotoxigenic *E. coli, C. jejuni*, enteroinvasive *E. coli/Shigella* spp., *Salmonella* spp., norovirus, and *Cryptosporidium* spp. An estimated 25%, 76%, and 0% of samples containing positive pathogen detects were accompanied by culturable *E. coli* in La Paz, Kanpur, and Atlanta, respectively, suggesting potential for viability of enteric microbes at the point of sampling. Airborne transmission of enteric pathogens merits further investigation in cities with poor sanitation.

**SYNOPSIS:** We detected and quantified molecular targets associated with important enteric pathogens in outdoor aerosols in cities with poor sanitation to assess the potential role of the aeromicrobiological pathway in enteric infection transmission in such settings.

## INTRODUCTION

With few exceptions, large cities in low- and middle-income countries (LMICs) have inadequate sanitation infrastructure^1–3^. Unsafe water and sanitation enable the transmission of enteric pathogens from infected individuals to susceptible hosts via direct contact or through the environment in multiple interconnected pathways^4,5^. While a rich and rapidly growing body of literature describes microbial risks associated with direct or indirect exposure to fecal contamination in a wide variety of settings, relatively few studies have examined the potential for transmission of enteric pathogens via the aeromicrobiological pathway in cities in LMICs. In these settings, the transport of enteric pathogens in aerosols may be possible due to a confluence of factors: inadequate sanitation infrastructure resulting in concentrated flows of fecal wastes, a high disease burden resulting in high-risk waste containing human enteric pathogens, high population density, and environmental conditions that may be conducive to the aerosolization of concentrated fecal wastes. The aerosolization, transport, and deposition of microbial pathogens in cities lacking good sanitation could lead to exposure either through inhalation or through ingestion via other pathways (e.g., food, water, direct contact)^6^. Aerosolization of biological material is known to be possible via several mechanisms including bubble bursting^7–9^, evaporation, raindrop impaction^10,11^, and others^12–15^. The creation and persistence of bioaerosols can be associated with a range of variables related to environmental conditions and the built environment including rain events^16–19^, meteorological conditions^20–22^, urban surface waters and water features^6,23,24^, wastewater treatment unit processes that include mechanical mechanisms^25,26^, and other infrastructure. The mechanisms behind aerosolization and transport of microorganisms from liquid surfaces and the microbial effects on droplet lifetime have been well-characterized under controlled conditions^7,8^. Laboratory studies have revealed that bubbles in contaminated water surfaces may experience conditions manipulated by microorganisms, allowing for smaller, more numerous, and higher velocity droplets to transition from water to air^9^. It has been shown that pathogens can be released via raindrop impaction, allowing for aerosol transport^10,11^. Despite an increasingly nuanced understanding of the physics of microbial transmission via aerosols, less is known about viability and persistence of infectious pathogens in this medium.

Among important human pathogens transmitted in aerosols, respiratory viruses including SARS-CoV-2 and influenza are the best characterized, primarily in indoor settings, given the central role of inhalation in exposures^27–31^. In the case of sanitation-related pathogens – where primary exposure is generally ingestion rather than inhalation – studies in high-risk, extramural (outdoor) settings in the USA and in other high-income countries have revealed that bioaerosols containing enteric microbes are common near point sources of concentrated fecal waste. Studies have primarily focused on ambient air surrounding wastewater treatment plants^32–43^ and in the context of land application of biosolids^44–54^; several studies have examined bioaerosols surrounding composting facilities^55,56^, meat markets^57^, impacted urban sites^58– 63^, and concentrated animal feeding operations^64–66^.

Of the studies examining bioaerosols at extramural urban sites, a majority have included a limited range of enteric microbes, generally reporting fecal indicator bacteria including members of the coliform group if using culture methods^60,67,68^, or reporting wastewater-associated or fecal genera typically from 16S sequencing data69–73 with rare population classification to the species or strain level. Targeted measurement of specific enteric pathogens is rare, partly because the presence of important enteric pathogens is unexpected outside settings where these infections are common, and few studies have been conducted where the burden of sanitation-related infections is high. While sequencing provides key information about airborne microbial communities by revealing relative abundance, quantitative estimates for specific pathogens are needed in exposure assessment and transport modeling^74^ and to further assess the potential public health relevance of this poorly understood pathway of transmission. Among previously reported studies of ambient urban aerosols, few have been located in LMICs despite the fact that cities in these countries are generally characterized by inadequate sanitation infrastructure and high population density. These conditions lead to widespread fecal contamination in close proximity to people, contributing to a high burden of disease. Diarrheal diseases caused an estimated 1.8 million deaths worldwide in 2017^75^, with LMICs bearing a disproportionate burden of morbidity and mortality.

Based on previous literature on the presence of enteric microbes in aerosols from well-studied settings in wealthy countries, we hypothesized that aerosolized enteric pathogens could be present and quantifiable where urban sanitation is lacking. We assessed this hypothesis in two cities with poor sanitation and in one city with established and maintained wastewater infrastructure as a reference site.

## METHODS

### Sampling locations

We conducted sampling in Kanpur, India (May – July 2017); La Paz, Bolivia (March 2018, June 2018, March 2019, June and July 2019); and Atlanta, Georgia, USA (March 2018-January 2019). Kanpur has distinct dry (October to June) and rainy (July to September) seasons; we sampled from May to August to capture both periods. Similarly, we intentionally sampled in La Paz during both rainy (December to March) and dry (May to August) seasons.

Kanpur is densely populated (Nagar district: 4.6 million people, population density of 1500 persons/km^2^)^76^ with a majority of untreated industrial, agricultural, and sewage waste conveyed via a system of uncovered canals (open wastewater canals, OWCs) discharging to the Ganges River^77,78^. In La Paz, a network of rivers receive untreated sewage discharge, industry effluent, and stormwater runoff; most of the waterway flows in a series of engineered channels^79,80^, also characterized as OWCs. The largest of these is the highly impacted Choqueyapu River, flowing through central La Paz (population: 900,000, 900 persons/km^2^)^81,82^ where it is joined by tangential tributaries including the Orkojahuira, Irpavi, and Achumani rivers. In past studies, this river system and its basin – eventually flowing into the Amazon – has been shown to contain a diverse and rich array of enteric microbes indicating high levels of fecal contamination^79,82,83^. As a reference site, Atlanta is characterized by having an established and maintained subsurface wastewater infrastructure, although urban surface waters in Atlanta’s watershed experience elevated levels of fecal-associated pathogens^84–86^ due to nonpoint source pollution and combined sewer overflows^87,88^. The city of Atlanta’s population density is an estimated 1500 persons/km^2^ ^89^, though sampling locations near impacted streams were in suburban locations at lower than mean population density.

We identified 18 sites in Kanpur, 37 sites in La Paz, and 8 sites in Atlanta meeting the following criteria: (1) proximity to sources of bioaerosols (<1 km) containing enteric microbes, OWCs in the cases of India and Bolivia and impacted surface waters in Atlanta; (2) public and ground level accessibility; and (3) unintrusive to members of the community during multi-hour sampling events. In Kanpur, we selected a control site greater than 1 km away from known OWCs and located on the Indian Institute of Technology IIT-Kanpur’s campus. The campus is a controlled private area with limited access to non-students and non-faculty, is less densely populated, has underground piped sewerage, and has a much lower animal presence. In La Paz, we identified two control sites >1 km from known concentrated wastewaters or other contaminated sources: (1) Chacaltaya, a weather station and environmental observatory located at 5380 m in elevation and far from human habitation and (2) Pampalarama, an unimpacted site near the Choqueyapu headwaters in a protected natural area. In Atlanta, we sampled at eight sites adjacent to impacted streams and rivers in Atlanta’s watershed: the Chattahoochee River, Proctor Creek, Foe Killer Creek, and South Fork Peachtree Creek. Additionally, we sampled on the roof of our laboratory and at ground level on Georgia Tech’s campus (located in Midtown Atlanta), both >1 km from surface waters as controls.

### Bioaerosol sampling, extraction and analysis

We used a combination of high-volume filtration and aerosol impaction in sampling across sites. We used the ACD-200 BobCat Dry Filter Continuous Air Sampler (InnovaPrep, Drexel, MO, USA) with sterile and single-use 52 mm electret filters and a flow rate of 200 L/min for downstream molecular analysis post extraction. We applied a sterile single-use wet foam carbon compressed elution kit (InnovaPrep, Drexel, MO, USA) to flush the filter following the manufacturer’s instructions, yielding approximately 6 mL of liquid eluate^90^. The pre-sterile single use filter and elution kit is intended to prevent cross-contamination between sequential samples; all filter handling and elution was conducted aseptically via manufacturer instructions and the high volume filter was wiped down with 10% bleach and 70% ethanol before and after sampling. As a control assessment for any residual background signal in the elution buffer and filters, we applied the eluate to the filters in the laboratory, extracted the blank eluate, and used this as a template for limit of detection calculations as well as sample blank controls in molecular analyses.

We treated the eluate with guanidine thiocyanate-based universal extraction (UNEX; Microbiologics, St. Cloud, MN, USA) lysis buffer in a 1:1 ratio, storing the mix in cryovials for sample transport to the laboratory. Samples were stored in a −80°C freezer immediately after transport to the laboratory, avoiding freeze-thaw by distributing the sample and buffer mixture into multiple aliquots. As a process control prior to extraction, we spiked the mix with 5uL of Inforce 3 Bovine Vaccine (Zoetis, Parsippany, NJ) containing bovine respiratory syncytial virus (BRSV) and bovine herpes virus (BoHV). Briefly, we conducted manual DNA and RNA extraction with the following steps: 1) added 300 uL of the mixture and 300 uL of 70% ethanol to a HiBind mini column (Omega BioTek, Norcross, GA), 2) following manufacturer advice to increase DNA yield, repeated step (1) three times, using a total of 450 uL of sample eluate, 3) washed the filter column with 100% ethanol, 4) and finally, eluted the nucleic acids and stored them in 50-75 μL of 10 mM Tris-1 mM EDTA (pH 8) at −80°C until further analysis^91^. Previous experiments have demonstrated an optimized recovery efficiency for *Cryptosporidium parvum* of around 50%^92^ and comparable crossing threshold (CT) values to other commercially available kits when assessed using real-time PCR^91^. In total, we collected 75 high-volume air samples from La Paz (71 collected near OWCs and 4 collected from control sites >1 km from OWCs), 53 high-volume air samples from Kanpur (45 collected near OWCs and 8 collected from one control site >1 km from OWCs), and 15 high-volume air samples in Atlanta (10 collected near impacted surface waters and 5 total (4 roof and 1 ground level) collected at the control site >1 km from impacted sites). We detail the time of day of sampling grouped by morning samples (AM, within the time period of 7am-12pm) and afternoon samples (PM, within the time period of 12pm-7pm) in addition to relevant information as described in the EMMI MIQE guidelines in Supporting Information^93,94^. The average volume sampled with the high-volume sampler was 47.5 m^3^ in La Paz (95% confidence interval = 42.3, 52.8), 36.3 m^3^ in Kanpur (95% confidence interval = 35.8, 36.7), and 28.3 in Atlanta (95% confidence interval = 30.4, 26.2).

For all high-volume air samples in Kanpur (n=53), we applied 1 mL of BobCat eluate to Compact Dry-EC (CD-EC) plates (Hardy Diagnostics, Santa Maria, CA, USA)^95^ for culture of total coliform and *E. coli*. Concurrent with high-volume air sampling in La Paz (n=31) and in Atlanta (n=15), we simultaneously used the Six-Stage Viable Andersen Cascade Impactor (ACI) with plates in six partitioned chambers at a flow rate of approximately 28.5 L/min for 1 hour to collect size-resolved bioaerosols in the size range of 0.65 to >7 μm (ACI, Thermo Scientific™, USA)^96^. We used AquaTest medium (Sisco Research Laboratories PVT. LTD., India) in the ACI to detect *E. coli*^97–99^. All culture samples were incubated at 37 °C and counted per the manufacturer’s instructions after 18-24 hours for colony forming units (CFUs). Due to poor potential recovery of culturable *E. coli* in high-volume samples from Kanpur, we subsequently used the more sensitive ACI in later sampling in La Paz and Atlanta. The average volume sampled in La Paz and Atlanta with the ACI was 1.30 m^3^ (95% confidence interval = 1.23, 1.37) and 0.970 m^3^ (95% confidence interval = 0.930, 1.01) respectively.

### Meteorological data

In La Paz, we collected solar UV irradiance (UVB, 280–320 nm), temperature (°C), and relative humidity (%) data from a stationary radiometer (Yankee Environmental Systems, Turners Falls, MA, USA) located approximately 2-5 km from sampling sites depending on location. In Kanpur, we collected on site data for each sample. We collected temperature (°C), and relative humidity (%) using the Vaisala HUMICAP Humidity and Temperature Transmitter Series HMT330 (Helsinki, Finland) and windspeed and direction with an anemometer (Ambient Weather, Arizona, USA). We conducted linear regression analyses to determine meteorological effects on enteric pathogens in aerosols. All analysis were conducted in R version 4.0.2^100^. We assessed the samples on a daily level as well as disaggregated the data by whether samples were taken in the morning (8am-12pm) or in the afternoon (12 pm and later).

### Enteric pathogen screening: multiplex qPCR

As a first step in screening enteric targets, we analyzed a subset of 40 high-volume samples from Kanpur and 23 high-volume samples from La Paz all <1 km from OWCs and 13 high-volume samples from Atlanta (7 of which were < 1 km from impacted surface waters) using a custom multiplex qPCR-based TaqMan Array Card (TAC) assembled and optimized by Thermo Fisher Scientific (Waltham, MA, USA). The subset was based upon the timing of the analysis performed and a limited stock of reagents. Select targets included globally important sanitation-related viruses (pan-adenovirus, pan-astrovirus, pan-enterovirus, norovirus GI/II, rotavirus A-C, and sapovirus I/II/IV/V), bacteria (*Aeromonas* spp., *Campylobacter coli, Clostridium difficile*, 9 *Escherichia coli* virulence genes (Table S1), *Enterococcus faecalis, Enterococcus faecium, Mycobacterium tuberculosis, Salmonella* spp., *Vibrio cholerae, Yersinia* spp.), protozoa (*Cryptosporidium parvum, Entamoeba histolytica, Giardia duodenalis*), and helminths (*Trichuris trichiura, Ascaris lumbricoides*) as well as multiple internal controls Though many of the targets are pathogenic, the card also includes some that may be commensal or of uncertain health relevance^101^, particularly in settings where asymptomatic carriage of common enteric pathogens is common^102^. We detail methods, descriptive statistics, targets, specific classifications of strains and types included in these assays, and their pathogenic relevance in the SI and Table S1, also previously described by Capone et al ^103^ and in accordance with the MIQE guidelines^104^.

### Quantitative molecular assays: ddPCR

For density estimation, we conducted absolute quantification of 12 enteric pathogen targets in high-volume aerosol samples via Droplet Digital PCR (ddPCR; QX200 Droplet Digital PCR System, Bio-Rad, Hercules, CA, USA). Targets included nucleic acids associated with selected viruses (adenovirus A-F, pan-enterovirus, norovirus GI, and norovirus GII), bacteria (*Campylobacter jejuni*, enteroinvasive *E. coli* (EIEC)/*Shigella*/spp., heat stabile enterotoxigenic *E. coli* (ST-ETEC), and two targets for *Salmonella* spp.), and protozoa (*Cryptosporidium* spp. and *Giardia duodenalis assemblage B*) and are detailed in Table S2. Targets represent a subset from TAC chosen based on public health relevance in the context of the global enteric disease burden^102^. Although by necessity a sub-set of globally important diarrheagenic pathogens, it includes those responsible for the highest burdens of morbidity and mortality resulting from acute diarrhea^105^ and that have been implicated in large-scale studies of diarrheal etiology^102,106–109^. We experimentally determined LODs for each assay using a probit analysis outlined by Stokdyk et. al^110^. A complete and detailed description of methods,pathogen targets, assay conditions, and all digital MIQE^111^ requirements are included in the SI. All data and metadata are accessible in a publicly accessible data repository (https://osf.io/NP5M9/)^93^

In addition to stratifying by sampling city and season (for La Paz and Kanpur), we further disaggregated data by distance from nearby putative sources. We collected GPS coordinates for each sample site and estimated the linear distance from the nearest OWC or impacted surface water. We use *a priori*-defined categories of 0-10 m and more than 10 m from OWCs (or impacted surface waters in Atlanta) and assessed the number of unique target detections per total assays we ran on each sample at each distance category. We used a Wilcoxon rank sum test to evaluate whether molecular detections decreased as a function of increasing distance from OWCs, time of day, and season. We used multiple linear regression to assess the effects of meteorological variables on target densities. All analyses were completed in R version 4.0.2^100^ and significance was based on 95% confidence (α=0.05).

## RESULTS

### Culturable fecal indicator bacteria (FIB) in aerosols

Because we could not measure pathogen viability directly across all targets at the point of sampling, we used culturable FIB as a proxy measure to indicate potential for viability of enteric microbes. Of the 45 air samples we collected in Kanpur near OWCs (<1 km) and analyzed by culture, 61% had detectable *E. coli* with an average concentration and 95% CI of 1.5 ± 1.3 CFU/m^3^_air_ across positive detections. Across all 45 samples including non-detects, the average concentration of *E. coli* was 0.92 ± 0.41 CFU/m^3^_air_. All control samples (n=7) taken >1 km away from observed fecal contamination were negative for culturable *E. coli*. In La Paz, adjacent to the Choqueyapu River and its adjoining tributaries, of the 28 air samples in close proximity to uncontained waste (<1 km) and analyzed for viable coliform, 52% were positive for *E. coli* with an average concentration and 95% CI of 11 ± 3.8 CFU/m^3^_air_. Across all 28 samples including non-detects, the average concentration of *E. coli* was 5.3 ± 2.1 CFU/m^3^_air_. We did not detect any *E. coli* in the samples taken >1 km away from uncontained waste (n=4). The size distribution capabilities of the ACI revealed that 27% of culturable *E. coli* were under 2.1 µm, the size cutoff for fine aerosol particles^112^. No samples in Atlanta were positive for *E. coli* though we detected total coliform (Figure 2).

**Figure 1.**
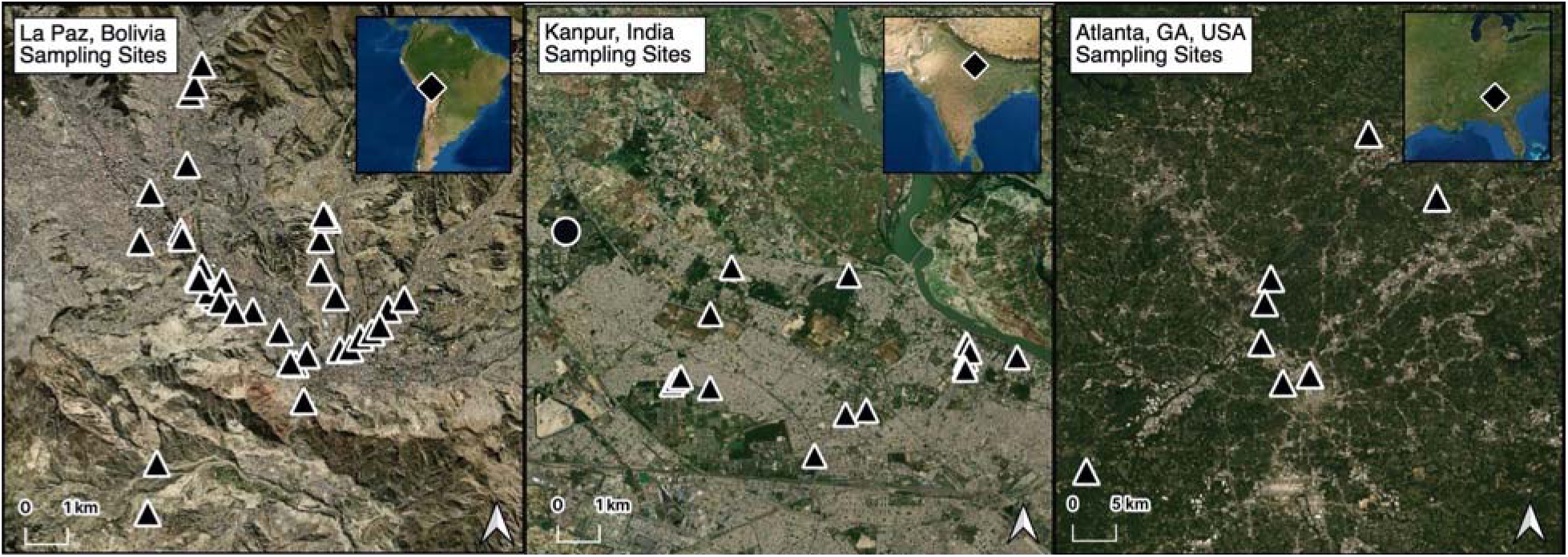
Aerosol sampling sites in La Paz, Bolivia; Kanpur, India; and Atlanta, USA. Sites located <1 km from open waste canals (OWCs) are represented by triangles and sites located >1 km from OWCs are represented by circles. Control sites (>1 km from OWCs) outside the city of La Paz not shown within this map extent but are located outside of the city of La Paz in remote areas.

**Figure 2.**
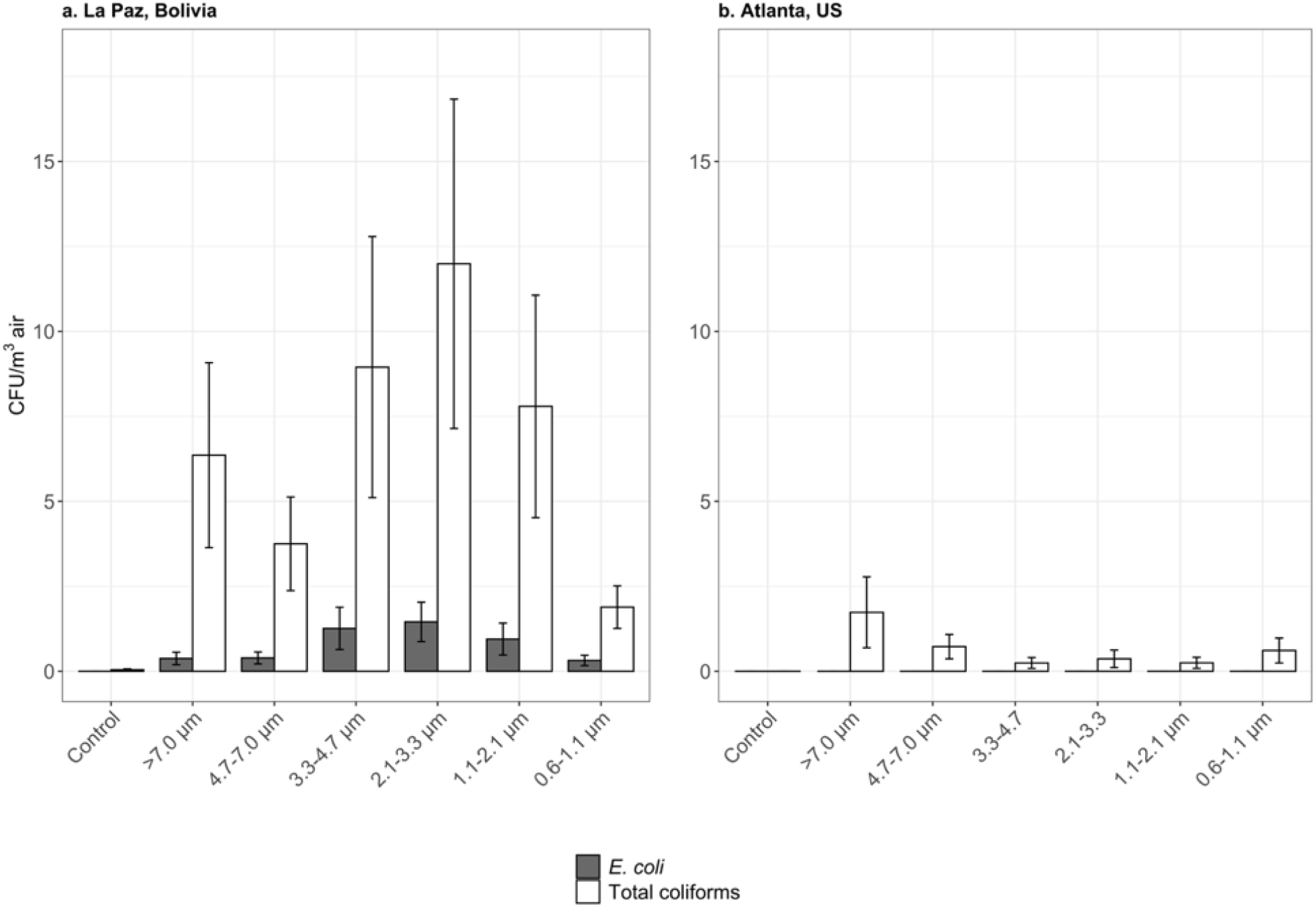
Size distribution of culturable *E. coli* and total coliform in La Paz and Atlanta. The mean standard error bars show 95% confidence intervals for densities in each size range. We did not collect Andersen Cascade Impactor (ACI) samples in Kanpur, so size-resolved *E. coli* detections by culture are not available for that site.

### Screening of enteric microbes in aerosols

We analyzed a subset of 40 high-volume samples from Kanpur, 23 high-volume samples from La Paz, and 13 high-volume samples from Atlanta for the presence and absence of 42 molecular targets including those specific to an *a priori*-defined list of globally important enteric viruses, bacteria, and protozoa. To compare across cities where we collected different numbers of samples, we assessed detections in the context of total volume collected at each site, normalizing to 2000 m^3^ for direct comparison. In La Paz, we detected genes associated with astrovirus and *Aeromonas* spp. in one of the three control samples. In Kanpur, we detected one each of the following molecular targets in control samples (n=8): adenovirus, non-typable rotavirus, *Aeromonas* spp., EIEC*/Shigella* spp., and *Yersinia* spp. At the control site in Atlanta (n=6), we detected *Aeromonas* spp. in one sample.

Positive detections for *a priori*-defined enteric pathogen-associated gene targets were noteworthy in samples taken less than 1 km from known fecal waste flows in Kanpur and La Paz. In Kanpur, 53% of these samples (n=13) were positive for at least one target, 28% (n=10) were positive for at least two targets, and 3% (n=4) were positive for at least five targets. Among these positive detections were genes associated with two protozoan parasites (*Cryptosporidium parvum* and *Giardia duodenalis*), four viruses (pan-astrovirus, pan-enterovirus, norovirus GII, and rotavirus), and five bacteria (*Aeromonas* spp., *Campylobacter coli*, pathogenic enteroaggregative *E. coli* (EAEC), *Enterococcus faecium*, and *Vibrio cholerae)*. In La Paz, 76% of samples less than 1 km from known fecal waste flows (n=16) were positive for at least one target, 62% (n=13) were positive for at least two targets, and 19% (n=4) were positive for at least five targets. Among these positive detections were five viral targets (adenovirus 40/41, pan-adenovirus, pan-astrovirus, pan-enterovirus, and norovirus GII), and nine bacterial targets (*Aeromonas* spp., EAEC, ST-ETEC, heat labile (LT)-ETEC, EIEC*/Shigella* spp., *Enterococcus faecium, Salmonella* spp., and *Yersinia* spp.). In Atlanta, 6 of 13 total samples were positive for one target (46%). Targets detected included one virus (pan-adenovirus) and two bacteria (*Aeromonas* spp. and *Campylobacter coli*). There were two Atlanta samples adjacent to the Chattahoochee River positive for adenovirus nucleic acid and one adjacent to Proctor Creek with *Campylobacter coli* nucleic acid, both above their respective 95% LODs (Table S2). The Chattahoochee commonly experiences levels of fecal indicator bacteria beyond EPA-recommended limits for recreational use, above 235 CFU per 100 mL^86,113,114^. To compare across cities, we normalized the collection volume to 2000 m^3^ and calculated the expected number of positives based on measured values. We also highlight the fraction of samples with positive detects that also tested positive for viable *E. coli* through culture (Figure 3). An estimated 25%, 76%, and 0% of samples containing positive pathogen detects were accompanied by culturable *E. coli* in La Paz, Kanpur, and Atlanta, respectively. We detected the highest pathogen diversity in La Paz, with genes specific to two enteric protozoa, ten bacteria, and six viruses among pre-defined targets. In La Paz, we detected multiple genes associated with a diverse array of pathogenic *E. coli* including EIEC/*Shigella*, LT and ST-ETEC, and two gene targets for EAEC. In Kanpur, we detected genes associated with two protozoa, five bacteria, and four viruses among pre-defined targets. In Atlanta, we detected gene targets associated with two bacteria and one virus with no co-detection of culturable *E. coli*. A linear regression model revealed a positive correlation between positive qPCR detects and viable *E. coli* detects in La Paz (p=0.02).

**Figure 3.**
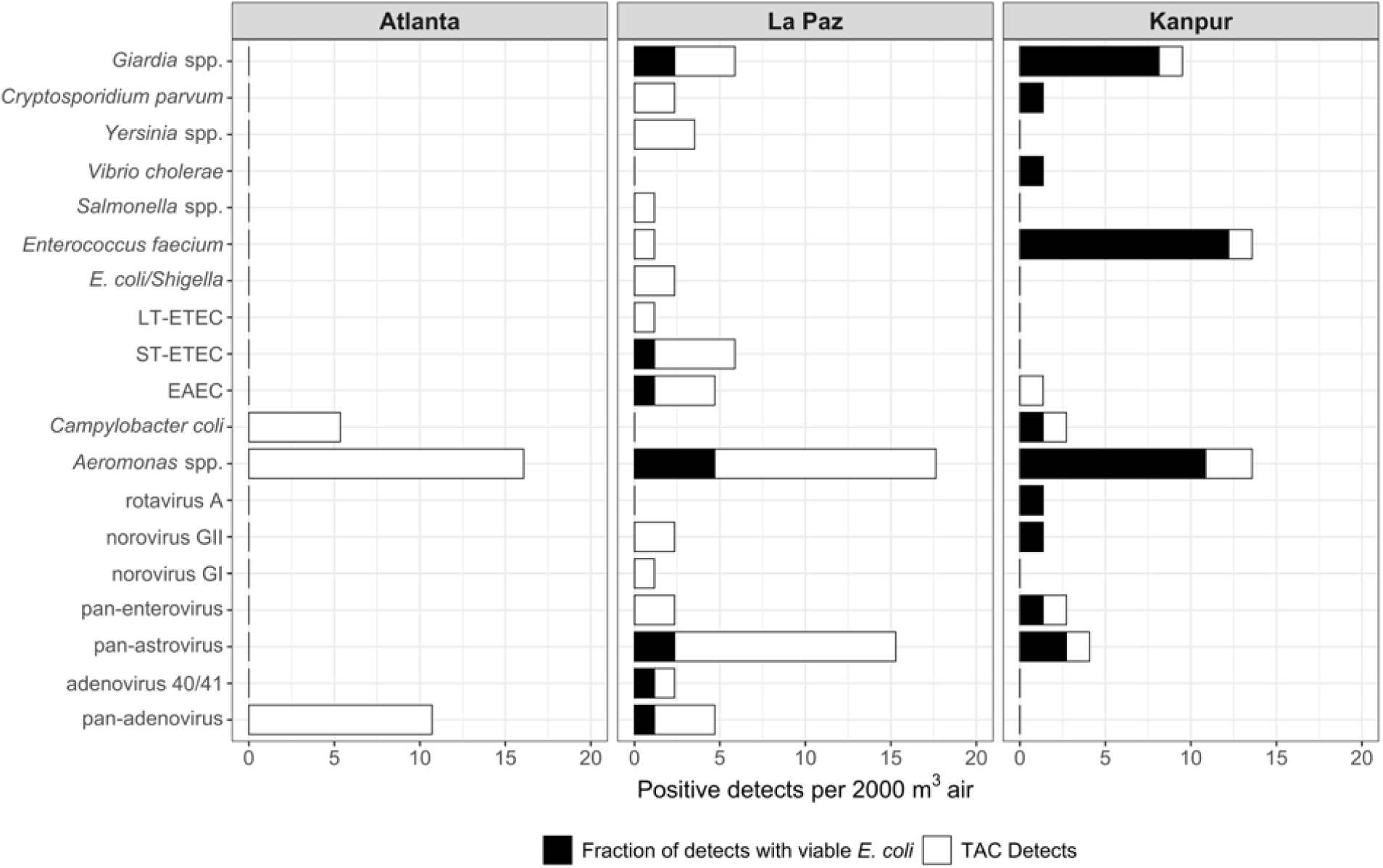
Positive detects via qPCR per 2000 m3 air and the fraction of positive detects with co-detection of culturable *E. coli* in the same sample.

### Quantitative estimation of enteric microbes in aerosols

We analyzed high-volume samples from the three sites for quantitative estimation of select, pre-defined gene targets associated with globally important enteric pathogens. We censored raw data using the 95% LOD as a conservative threshold for positivity. In Atlanta, we quantified *Shigella* spp.*/*EIEC (3 detections). In La Paz, we quantified ST-ETEC (2 detections), *Shigella* spp.*/*EIEC (16 detections), *Campylobacter jejuni* (2 detections), *Salmonella* spp. ttr gene (3 detections), pan-enterovirus (3 detections), adenovirus A-F (1 detection), norovirus GI (3 detections), norovirus GII (3 detections), and *Cryptosporidium* spp. (3 detections). In Kanpur, we quantified *Campylobacter jejuni* (1 detection in 53), norovirus GI (33 detections), norovirus GII (1 detection), MS2 (1 detection), and *Cryptosporidium* spp. (3 detections). Among the detections above the 95% LOD threshold, target densities ranged from 1.5×10^1^ gc per m^3^_air_ to 4.7×10^2^ gc per m^3^_air_ (Figure 4).

**Figure 4.**
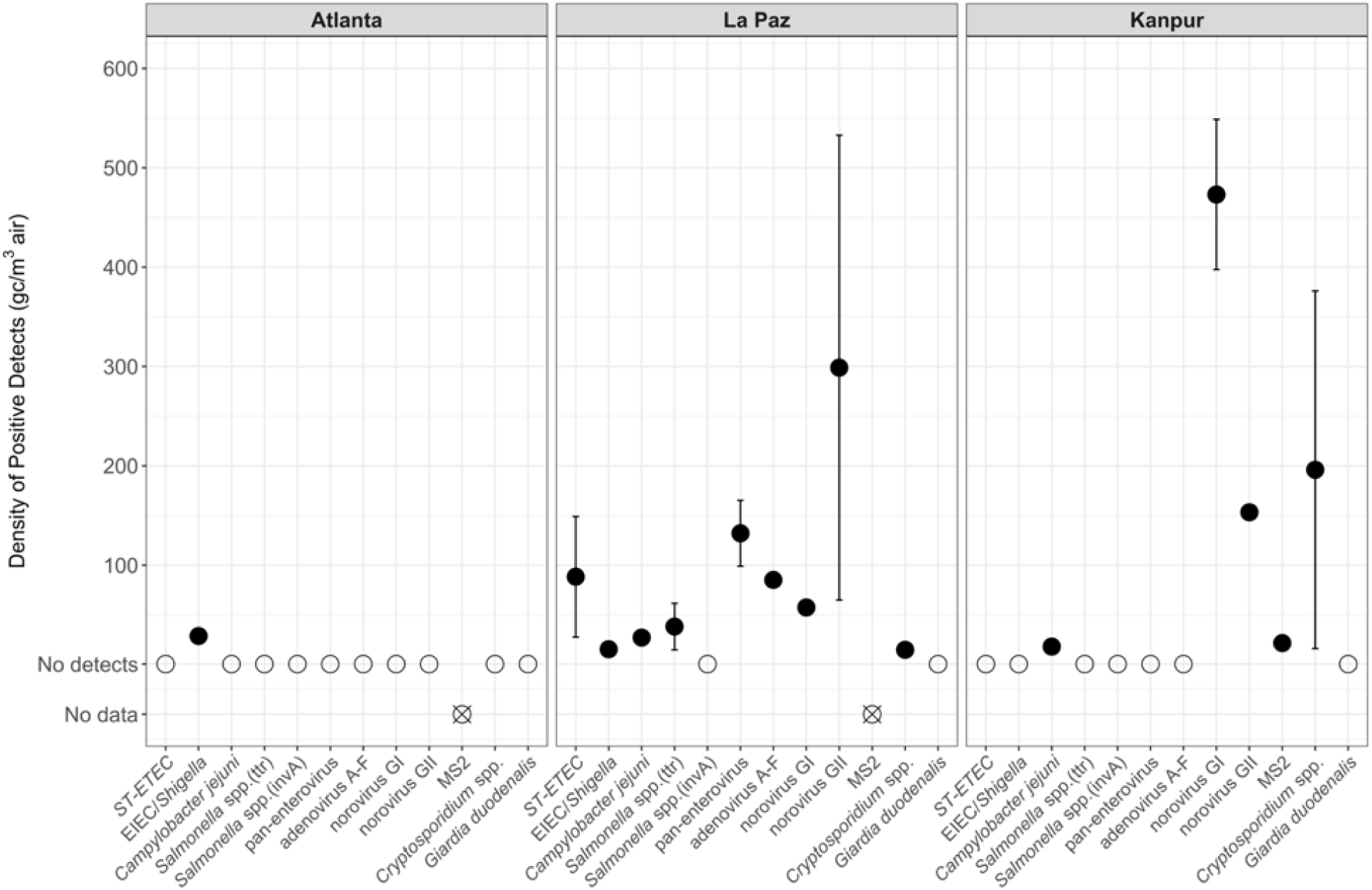
Mean densities of gene targets associated with enteric microbes with mean standard error bar as observed among the distribution of positive detects in gene copies per cubic meter of air. Densitie were censored according to the assay-specific 95% limit of detection (LOD)

We observed some differences in detection of enteric targets (gene copy density estimates) across meteorological variables, though interpretation of these associations is constrained by limited sample size. In La Paz, temperature was the only variable we measured that had a statistically significant effect on target density. In a stratified analysis from morning sample times only, our linear regression model showed that temperature was the only meteorological variable with any effect on target densities. In assessing the direct relationship between temperature and each target, we found that as temperature increased, pan-adenovirus, ST-ETEC, EIEC/*Shigella spp*., norovirus GI and norovirus GII density decreased (p=0.004, p=0.01, p=0.02, p=0.03, and p=0.01 respectively). We detected no significant effect of any variables on target densities later in the day. Ungrouped by time of day and with analysis restricted to average daily meteorological measurements, among all samples we detected no statistically significant effects of any variables we examined on pathogen target densities. When grouped by season in La Paz (rainy or dry), we found that *E. coli* density increased in the rainy season (p=0.05), on average. Mean temperature, relative humidity, and UV were higher in the rainy season. In Kanpur, we observed higher norovirus GI and GII densities in the dry season, before the monsoon (p=0.00003 and p=0.05 respectively). Grouped only by time of day and not season, we detected norovirus GI at greater densities in the morning compared to samples taken in the afternoon (p=0.03). In assessing the effects of meteorological measurements on target densities using a multivariable linear model, we found relative humidity to be the only variable with any effect on target densities. As relative humidity as measured by Vaisala increased, norovirus GI and GII density decreased (p=0.04 and p=0.03). A complete accounting of all relevant model results is included in the SI (Table S3).

We stratified data by distance from OWCs (for Kanpur and La Paz) or impacted urban water sources (Atlanta) (Figure 5). The number of unique targets detected decreased as distance from OWCs increased in La Paz and Kanpur. We observed a downward trend in density for one target in La Paz (EIEC/*Shigella* spp.) and Kanpur (norovirus GI). Overall, we observed a clear decrease (p=0.005) in the probability of detecting any positive molecular pathogen target (including ddPCR and qPCR detections above LOD) between samples collected within 10 m and samples collected greater than 10 m from OWCs in La Paz and Kanpur and impacted urban surface waters in Atlanta. We also observed a decrease in the probability of detecting bacterial, viral, and protozoan molecular pathogen targets (p=0.006, p=0.02, and p=0.009 respectively) between samples collected within 10 m and samples collected greater than 10 m from potential sources. We observed no differences in probability of detection across all targets by seasonality (rainy/dry) in La Paz and Kanpur.

**Figure 5.**
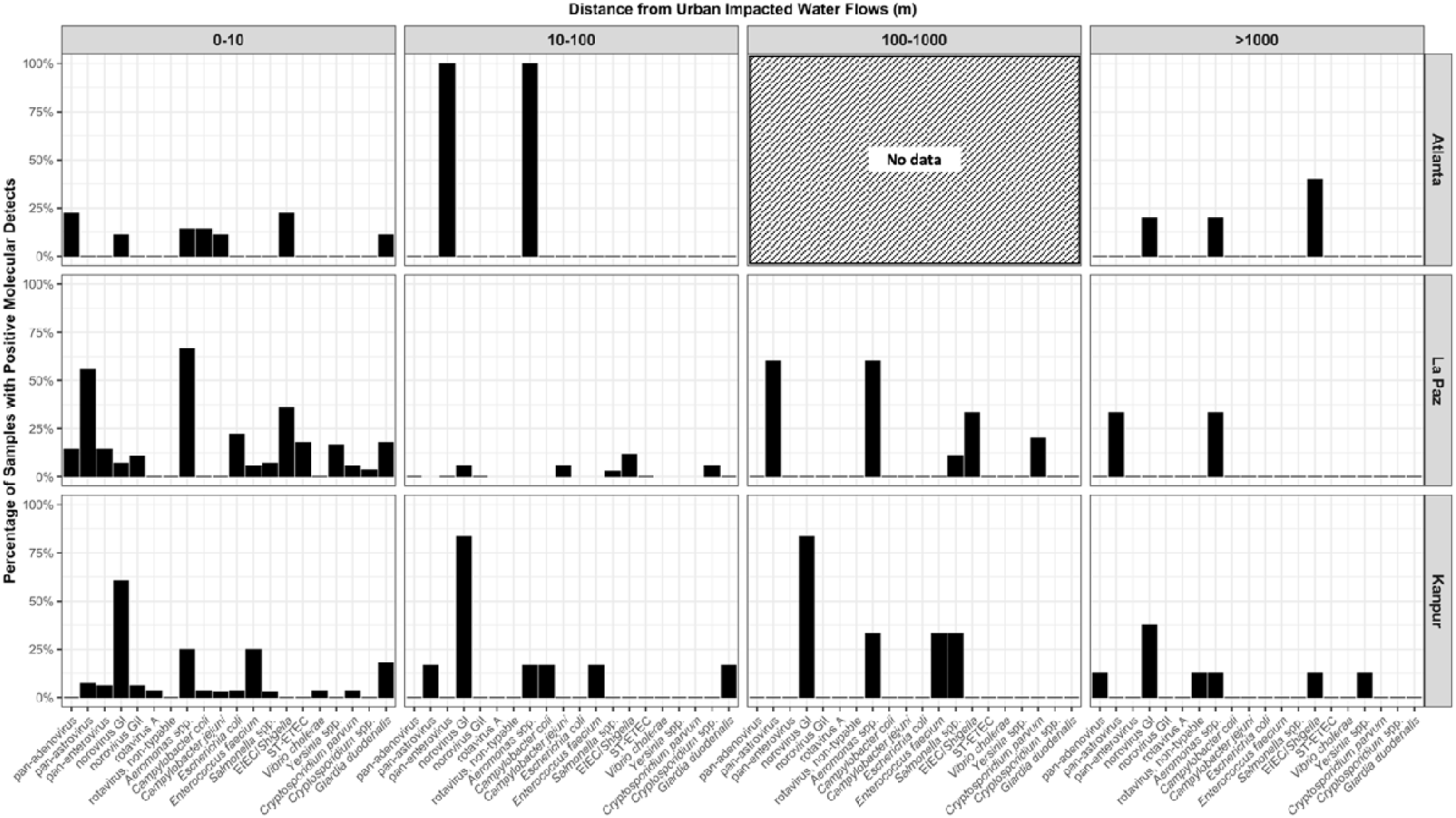
Percentage of positive detects (via qPCR or ddPCR) in sample sets for each site versus straight-line distance from impacted water flows, where densities were censored according to the assay-specific 95% limit of detection (LOD).

## DISCUSSION

Open sewers conveying domestic, institutional, commercial, and industrial effluent are common in cities in LMICs. They may also serve important drainage functions including flooding control. Sewers may be open to the atmosphere because solid waste can clog closed drains, and a lack of adequate solid waste management in some cities makes OWCs a rational approach to removing concentrated waste away from human habitation. They may pose risks, however, both to downstream communities and people in close proximity to open urban wastewater flows.

Our results suggest that in cities of LMICs with poor sanitation infrastructure and the presence of concentrated, uncontained fecal waste streams in open sewers, genes specific to enteric microbes, many pathogenic, are present in aerosols and may disseminate in the local environment. Overall, detection of fecal microbes in aerosols was higher than we expected at each of our study sites. The diversity and density of enteric microbes was enriched in La Paz and Kanpur compared with reference sampling in Atlanta and was greater near open wastewater canals and impacted surface waters compared to sites farther from known potential sources. The co-detection of culturable *E. coli* in a high percentage of samples in La Paz and in Kanpur suggests, indirectly, that some bacterial pathogens detected could have been viable at the point of sampling. We observed no culturable *E. coli* in aerosols sampling in Atlanta. The health risk implications of the presence of aerosolized enteric microbes in these settings are unknown but merit further study.

Our study included a range of pathogen targets of global public health relevance, many of which have not been previously detected in urban outdoor aerosols where infection risk is a clear possibility due to the proximity of concentrated waste and high population density. In La Paz, we quantified ST-ETEC in two aerosol samples at densities of 28 gc/m^3^_air_ and 150 gc/m^3^_air_. ETEC was responsible for 50,000 deaths worldwide in 2016^115^ but has not been previously quantified in extramural aerosols in cities where transmission is endemic. Also in La Paz, we report the first quantitative estimate of EIEC/*Shigella* (*ipaH* gene, n=16) in a similar setting at densities ranging from 1.8 gc/m^3^_air_ to 53 gc/m^3^_air_. We detected and quantified other enteric bacteria not previously observed in extramural urban aerosols such as *Campylobacter coli* and *Salmonella* spp., though they have been previously quantified in air near concentrated animal feeding operations^116,117^.

We observed comparable prevalence of *Aeromonas spp*. associated nucleic acids in Atlanta, La Paz, and Kanpur with 8, 9, and 7 positive detections per 2000 m^3^_air_ at each site respectively. *Aeromonas* spp. have been consistently detected in environmental media in a variety of settings^118^. Although some aeromonads are important human pathogens, in 2016 *Aeromonas* spp. were responsible for only 1% of total diarrheal deaths and only 19 of 36 subtypes are known to be pathogenic.^115,119^

Among viral detects in Kanpur and in La Paz, norovirus GI and GII may be most relevant to health. We detected norovirus GI and GII at the highest average density across all targets in Kanpur aerosol samples (320 gc/m^3^_air_ and 150 gc/m^3^_air_ respectively). In La Paz, we detected norovirus GII at the highest average density across all targets (13 gc/m^3^_air_) and norovirus GI at a mean of 2.4 gc/m^3^_air_. Norovirus is relatively resistant to inactivation in environmental media^120^, may persist on environmental surfaces for up to two weeks^121^, has been shown to survive in aerosols^122–126^, and has an estimated relatively low median infectious dose between 18 and 10^3^ virus particles^127^. It is among the most widely prevalent viral enteric pathogens, globally, with an estimated 33,000 cases and 20,000 deaths per year^115,128,129^. Norovirus transport through bioaerosols and subsequent deposition and indirect exposure through ingestion remains poorly characterized.

We are aware of only one previous study reporting detection of enteric protozoan parasites in air samples, from rural Mexico, by microscopy; the study reported 8 of 12 samples positive for *Cryptosporidium* and 10 of 12 samples positive for *Giardia*, possibly via aerosolization of soil^130^. By comparison we detected *Giardia duodenalis* via qPCR in 22% of samples in La Paz and 18% of samples in Kanpur, with 3 and 5 positive detections per 2000 m^3^_air_ at each site respectively. *Cryptosporidium* was present in 9% and 3% of samples in La Paz and Kanpur respectively with 1 positive detection per 2000 m^3^_air_ at both sites. We quantified *Cryptosporidium* spp. in aerosol samples via ddPCR in La Paz (n=3) and in India (n=2) at average densities ranging from 9.3 gc/m^3^_air_ to 560 gc/m^3^_air_, the second time *Cryptosporidium* has been reported in an aerosol and the first quantitative estimate. *Cryptosporidium* is among the most prevalent etiologies of moderate to severe diarrhea globally^102,131^.

A small number of previous studies have identified the likely presence of aerosolized fecal material and potential for pathogen transmission in bioaerosols in similar settings. Our previous studies in La Paz and Kanpur reported detection of culturable *Enterobacteriaceae* and *E. coli* in ambient urban air that was hypothesized to also originate from OWCs^132–134^. A study in Malawi measured the presence of enteric microbes in ambient air (enterotoxigenic *E. coli*) before, during, and after pit latrine emptying, confirming that these microbes increased in nearby aerosols during pit latrine emptying events^67^. In 22 samples of outdoor aerosols collected in Mumbai, 28 species of culturable bacteria were identified including several opportunistic pathogens: *Staphylococcus* spp., *Serratia plymuthica*, S*erratia haemolyticus*, and *Enterobacter aerogenes*^60^. Bacterial bioaerosols including opportunistic pathogens have similarly been identified using 16S rDNA sequencing near composting facilities in India^56^. *Staphylococcus aureus* and other opportunistic pathogens have been identified in urban environments in the Philippines^135^. A larger study from Beijing, China identified many of the same genera in addition to 16 species of *Pseudomonas* (some potential opportunistic pathogens) and the possibly fecal-associated genera *Enterococcus, Escherichia, Vibrio*, and *Yersinia*^59^.

The concurrent detection of culturable *E. coli* in many samples from La Paz and Kanpur suggests that some of these important pathogenic bacteria (including pathogenic *E. coli*), viruses, and protozoa we detected may have been viable at the point of sampling^136^. As a commonly used fecal indicator bacterium, *E. coli* suggests – though does not conclusively demonstrate – the presence of aerosolized fecal material in samples^137^. Culturable *E. coli* also may indicate recently aerosolized material, since vegetative bacteria are not persistent in the aerosolized state and may be quickly inactivated if ideal conditions are not met.^102^ Other sanitation-related pathogens, such as those showing greater persistence in the environment (e.g., *Cryptosporidium* oocysts) may persist for longer periods in environmental media^138^ than co-occurring vegetative bacteria^139^, suggesting that culturable *E. coli* may conservatively represent the potential viability of other enteric pathogens, particularly protozoa. We further note that we have used culturable *E. coli* as an indicator of viability only, rather than an indicator of exposure risk: no relevant standards for *E. coli* or other FIB in aerosols exist that would indicate a threshold of exposure risk in these settings. Also, differences in methods for *E. coli* capture and culture were different across sites preventing a direct quantitative comparison of the data to inform risk on the basis of *E. coli* counts. More work on the comparative survival of pathogens in extramural aerosols is needed. However, such studies are challenging to conduct, because the methods for capturing aerosolized enteric pathogens at low densities – requiring high flow-rates – are not ideal for preserving viability^140–142^.

Size-resolved densities of culturable *E. coli* indicate the presence of enteric bacteria in aerosols ranging from 0.6 µm to 7 µm. Culturable *E. coli* that were captured under 2.1 µm^112^ indicate possible suspension of bioaerosols over a period of hours, with a settling velocity in still air of 0.5 m/hour for a typical particle with a 2 µm diameter^143^, indicating high transport potential in air near OWCs. Larger diameters indicate that the fecal bacteria may deposit on surfaces more rapidly. The size-resolved density estimates of FIB may or may not indicate the size ranges associated with other enteric microbes, including viral and protozoan pathogens, but may be useful as an initial proxy for use in transport modeling for enteric microbes. Though more work is needed to characterize exposure potential based on aerosol size, and although some enteric pathogens have been known to cause respiratory infections^144–146^, the most likely pathways of exposure relevant to bioaerosols in this setting are associated with surface deposition of pathogens and subsequent transport via other well understood pathways represented in the F-diagram^5^. There is some epidemiological evidence that proximity to concentrated fecal waste streams in urban areas can be related to enteric infection risk. Contreras *et al*.^147^, assessing the spatial relationship between household proximity to fecal contaminated surface water canals used for crop irrigation and diarrheal disease in children in multiple municipalities in the Mezquital Valley in Mexico, determined that compared to children under 5 living within 10 m from a canal, children living 100 m from a canal had 45% lower odds of diarrhea and children living 1000 m from a canal had 70% lower odds of diarrhea. They further estimated that 24% of all diarrheal cases in the study and 50% of all cases within 100 m from a canal were attributable to canal exposure. The authors posited aerosolization of pathogens from canals as a potential pathway of exposure in this setting. Our results confirm the presence of enteric pathogens in aerosols at these distances and we report a statistically signficant decrease in bacterial, viral, and protozoan molecular detects as distance increases from a potential source. Though the density range of gene copies per m^3^ that we quantified in aerosols is lower than what we might find in concentrated sources such as wastewaters, microbial infection risks attributable to aerosols in these settings cannot be ruled out given co-detection of culturable FIB and close proximity to population centers. Important next steps include further epidemiological studies examining risks attributable to this pathway as well as mechanistic studies including quantitative microbial risk assessment. Such studies would be useful in estimating both direct and indirect infection risks that account for microbial transport and viability in aerosols and following deposition via other pathways of relevance in fecal-oral transmission, including comparative risk assessment that examines the relative contributions of multiple pathways of exposure.

Our quantitative estimates of specific pathogens are a necessary first step towards further work in understanding the implications of the presence of these microbes in air, including fate and transport modeling and risk assessment.

This study had a number of important limitations that deserve consideration. First, the viability of enteric pathogens detected by molecular methods in aerosols cannot be assumed, even with co-culture of fecal indicator bacteria. Although *E. coli* viability in samples may indirectly indicate potential viability of other microbes present in bioaerosols, and vegetative bacteria may represent a conservative proxy since they have been shown to survive relatively poorly in aerosols^138,148^, we did not measure viability of other microbes directly. The methods we used for high-volume samples present high pressure and desiccating conditions that may reduce viability of microbes captured on the filter^149^, and may also have resulted in an underestimation of culturable total coliforms and *E. coli* in Kanpur; in contrast, the ACI used in Bolivia and Atlanta is suitable for coliform survival and growth since bioaerosols are impacted at a lower flow rate onto nutrient-rich, semi-solid agar media. High-volume aerosol sampling generally presents conditions that are known to limit recovery of viable microbes^150^. Capture methods that preserve viability, such as impingement^65^ and water-vapor condensation^28^ typically operate at relatively low flow rates (8 - 13 liters per minute), requiring extended periods to capture targets present at low densities. We further acknowledge that pathogen-specific nucleic acids in aerosols could be attributable to either viable or inactivated microbes, or may exist as extracellular genetic material in the environment. Second, while our data suggest concentrated fecal waste streams as potential sources of aerosolized sanitation-related microbes in nearby air samples, and we observed a trend of decreasing density with increasing distance from fecal waste streams, we cannot definitively conclude that enteric microbes in aerosols derive from these sources. Cities with poor sanitation infrastructure typically have many contaminated sites, including OWCs and drains but also open latrine pits^67,151^, composting sites^56^, or uncontained solid waste^152,153^.

Animals and animal waste may be common and could be aerosolized. Further work is needed on methods for source-tracking of bioaerosols, including via sequencing approaches. Third, our molecular results are likely conservative representations of target densities in ambient air of the sampling locations based on laboratory experiments with the high-volume sampler that reveal recovery efficiencies of 1 µm particles ranging from 73% in conditions most similar to those of our study^150^ to 101% under controlled laboratory conditions^154^. Finally, this study included a limited sample size across the wide range of fecal-related targets we sought, constraining statistical power for assessment of risk factors for pathogen detection across sites. Further studies of specific pathogen transport under specific controlled conditions are needed to fully describe mechanisms of aerosolization, transport, deposition, viability and persistence in aerosols, and risk of exposure to humans.

We highlight aerosol transport as playing a potentially important and understudied role in the spread of microbes originating in fecal wastes in outdoor environments with uncertain implications for human exposure, infection, and disease transmission. Fecal-oral transmission of enteric pathogens is often summarized in the so-called F-diagram, describing key media that serve as direct and indirect sources of enteric pathogen spread: water, soil, hands, fomites and surfaces, food, and flies^5,155,156^. It is possible that aerosols should be added to this conceptual framework contingent on further work in this area. Aerosols may allow for transport of enteric pathogens between and among media, contributing to the spread of fecal contamination and associated microbes, resulting in potential for greater exposure via contact, inhalation, or ingestion either directly or indirectly following deposition on a surface, food, water or other subsequent exposure pathway^6,147^. In many settings, multiple relevant pathways of exposure may exist^157^, and aerosols may be one more whose risk relevance remains uncharacterized but cannot be excluded from consideration. There have been no previous attempts to estimate the global, national, or local burdens of morbidity and mortality attributable to airborne transmission of enteric pathogens in urban outdoor environments, partly because this phenomenon is poorly understood and has not been studied in either epidemiological research or risk assessment at scale. Indeed, our paper is the first to quantitatively describe the potential for transmission in settings where enteric pathogens are common and therefore represent potentially important exposure risks. We hope this work stimulates further work in epidemiology and infectious disease research to advance further understanding of this pathway and to estimate attributable burdens of disease and death, where appropriate. A complete accounting of enteric pathogen transport is required to design intervention strategies with the potential to control exposures. In high-burden settings such as rapidly densifying cities in LMICs with poor sanitation, enteric pathogen transport via aerosols near concentrated fecal waste flows merits further investigation. Direct measures of pathogen viability and persistence in aerosols, exposure modeling, quantitative microbial risk assessment, and epidemiological studies would be useful next steps in further characterizing the public health relevance of this phenomenon.

## Data Availability

All study data are available at Open Science Framework: https://osf.io/NP5M9/.

## ABBREVIATIONS

(LMIC): Low-and middle-income country
(OWC): open wastewater canal
(BRSV): Bovine respiratory syncytial virus
(BoHV): Bovine herpes virus
(TAC): Taqman Array Card
(ddPCR): Droplet digital PCR
(EIEC): Enteroinvasive *E. coli*
(ST-ETEC): Heat stabile enterotoxigenic *E. coli*
(EAEC): Enteroaggregative *E. coli*
(LT): Heat labile
(LOD): Limit of detection

## SUPPORTING INFORMATION

Tables: 3

Figures: 3

Pages: 21

## ACKNOWLEDGEMENTS

This material is based upon work supported by the National Science Foundation under grant number 1653226. Many thanks to colleagues at IIT-Kanpur including Hari Shankar, Harish Vishwakarma, Shivshankar Mishraa and numerous others who helped with instrumentation and shared their laboratory space. We further acknowledge the kind people who shared their homes and their tea with us as we sampled in the community. Thanks to all colleagues at Universidad Católica Boliviana and Universidad Mayor de San Andrés, including undergraduate assistants.

## TOC ART: FOR TABLE OF CONTENTS ONLY

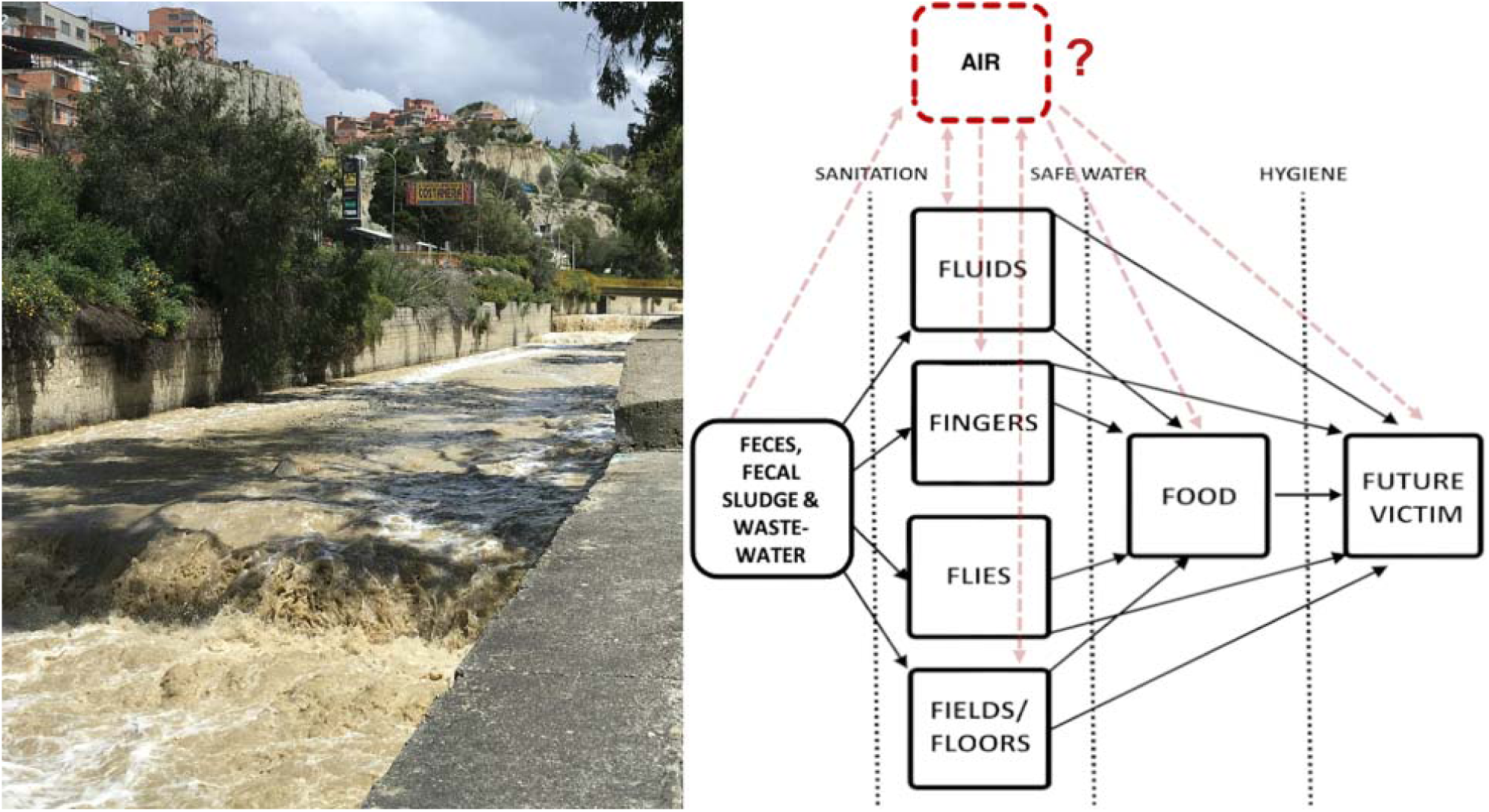

