## Supporting Information for "Detection and quantification of enteric pathogens in aerosols near open wastewater canals in cities with poor sanitation"

**Contents.**

Tables: 3

Figures: 3

Pages: 21

**Custom TaqMan Array Card (TAC).**

We purchased custom TACs assembled and optimized by Thermo Fisher Scientific (Waltham, MA, USA). The TAC is a 384-well array of μL reaction vessels with dried-down primers and hydrolysis probes for the defined targets for amplification using TaqMan real-time PCR technology. Each array card has 8 ports for loading samples. To prepare the samples for PCR, we mixed 50 μL of total nucleic acid template (0.5 μL template per reaction well) with 50 μL of qScript XLT 1-Step RT-qPCR ToughMix (Quantabio, Beverly, MA) and filled ports 2-7 with the combined 100 μL for each sample. Port 1 was used as a negative control and port 8 was used as a positive control, allowing us to include 6 samples per card. For the NTC we used molecular water extracted using the same protocol as the samples. For the PC template, we used a single-use aliquot mixture of nucleic acid for each target (gene targets inserted into plasmids) (IDT, Coralville, IA) which were developed using methods previously described^1^

Following the manufacturer’s instructions, we centrifuged each card twice at 1200 rpm for one minute, sealed the card, trimmed the loading ports and loaded the card into a QuantStudio 7 (Thermo Fisher Scientific, Waltham, MA). Reverse transcriptase real-time PCR was performed using the following cycling conditions with a 1°C/s ramp rate between all steps: 45°C for 10 minutes, 94°C for 10 minutes, and then 45 cycles of 94°C for 30 seconds and 60°C for 1 minute. All positive controls amplified as expected (average Ct = 27 across all assays) and we detected MS2 in all samples. Additionally, the TAC included an internal positive control (TaqMan^TM^ Exogenous Internal Positive Control, Applied Biosystems, Foster City, CA), which we used to monitor for potential inhibition. The internal positive control assay amplified consistently with no indication of inhibition for all air samples (average Ct = 24, range = 22-26). We observed positive amplification of our positive control for all assays (n=32). We observed no amplification for any assay in any no template controls (n=13) below a quantification cycle (Cq) of 40, the cutoff we used for positive detects as has been used previously^2,3^. The threshold of amplification was set for each individual assay at the point of inflection and we interpreted samples as positive if there was a clear distinction between the positive and negative amplification curves

**Table S1**. TAC targets, general description, further classification specificity, and pathogenicity.

| **Category** | **General description** | **Further classification** | **Interpretation** | **Assay reference** | **Generally pathogenic in immunocompetent hosts** |
| --- | --- | --- | --- | --- | --- |
| Bacteria | *Aeromonas* | *Aeromonas hydrophila, caviae, veronii, jandaei, salmonicida, schubertii, popofii* | If detected, call as *Aeromonas* spp. positive | ^4^ | no ^5^ |
|  | *Campylobacter coli* | *Campylobacter coli (cadf gene)* | If detected, call as *Campylobacter coli/jejuni* positive | ^6^ | yes^7^ |
|  | *Clostridium difficile* | *Clostridium difficile* toxin A gene tcdA (toxigenic *Clostridium difficile*) | If either detected, call as *Clostridium difficile* positive. | ^8^ | yes^9^ |
|  | *Clostridium difficile* | *Clostridium difficile* toxin B gene tcdB (toxigenic *Clostridium difficile*) |  | ^8^ | yes^9^ |
|  | *Clostridium difficile* | Non-toxigenic *Clostridium difficile* when a non-coding insertion sequence is present in the pathogenicity locus (PaLoc) | If detected, call as non-toxigenic *Clostridium difficile* positive | ^10^ | no^11^ |
|  | EAEC | Enteroaggregative *Escherichia coli* (aaiC gene) | If either was detected, call as EAEC positive | ^4^ | yes^12^ |
|  | EAEC | Enteroaggregative *Escherichia coli* (aatA gene) |  | ^13^ | yes^12^ |
|  | EIEC/*Shigella* | Enteroinvasive *Escherichia coli/Shigella* (ipaH gene) | If detected, call as *Shigella*/EIEC positive | ^14^ | yes^15^ |
|  | *Enterococcus* | *Enterococcus faecalis* | If detected, call as *E. faecalis* positive | ^10^ | no^16^ |
|  | *Enterococcus* | *Enterococcus faecium* | If detected, call as *E. faecium* positive | ^10^ | no^16^ |
|  | EPEC | Enteropathogenic *Escherichia coli* (eae gene) | If either was detected, call as EPEC positive | ^4^ | yes^17^ |
|  | EPEC | Enteropathogenic *Escherichia coli* (bfpA gene) |  | ^4^ | yes^18^ |
|  | LT-ETEC | *Escherichia coli* (heat-labile enterotoxin) | If detected, call as LT-ETEC positive | ^19^ | yes^20^ |
|  | ST-ETEC | *Escherichia coli* (heat-stable enterotoxin) | If detected, call as ST-ETEC positive | ^4^ | yes^20^ |
|  | *Salmonella* spp. | *Salmonella bongori* and all subspecies of *Salmonella enterica* | If detected, call as *Salmonella spp.* positive | ^4^ | yes^21^ |
|  | Shiga-like toxin 1 | shiga toxin carried by *Shigella dysenteriae*; shiga-like toxin 1 carried by *Escherichia, Citrobacter, Aeromonas*, or *Enterobacter* genus | If either was detected, call as STEC positive | ^4^ | yes^22^ |
|  | Shiga-like toxin 2 | shiga-like toxin 2 carried by *Escherichia, Citrobacter, Aeromonas*, or *Enterobacter* genus |  | ^19^ | yes^22^ |
|  | *Vibrio cholerae* | *Vibrio cholerae* with or without the cholera toxin-encoding gene | If detected, call as *Vibrio cholerae* positive | ^4^ | no^23^ |
|  | *Vibrio cholerae* | *Vibrio cholerae* carrying the cholera toxin-encoding gene |  | ^4,24^ | yes^25^ |
|  | *Yersinia spp.* | all species within the *Yersinia* genus | If detected, call as *Yersinia spp.* positive | ^26^ | yes^16^ |
| Viruses | adenovirus 40/41 | adenovirus serotypes 40 and 41 | If detected, call as adenovirus 40/41 positive | ^27^ | yes^28^ |
|  | pan-adenovirus | adenovirus serotypes except 40 and 41 | If detected, call as pan-adenovirus positive | ^10^ | no^29^ |
|  | pan-astrovirus | all human serotypes of astrovirus | If detected, call as pan-astrovirus | ^30^ | yes^31^ |
|  | pan-enterovirus | all enterovirus serotypes with the enterovirus genus | If detected, call as pan-enterovirus | ^24^ | no^32^ |
|  | norovirus GI | norovirus GI | If detected, call as norovirus GI positive | ^33^ | yes |
|  | norovirus GII | norovirus GII | If detected, call as norovirus GII positive | ^34^ | yes |
|  | rotavirus | rotavirus A | If detected, call as rotavirus A positive | ^35^ | yes |
|  | rotavirus | rotavirus B | If detected, call as rotavirus B positive | ^10^ | yes |
|  | rotavirus | rotavirus C | If detected, call as rotavirus C positive | ^10^ | yes |
|  | rotavirus | rotavirus non-typable | If detected, call as pan-rotavirus | ^10^ | yes |
|  | sapovirus V | sapovirus belonging to genogroup V | If either detected, call as Sapovirus positive | ^4^ | yes^36^ |
|  | sapovirus I, II, or IV | sapovirus belonging to genogroups I, II, or IV |  | ^4^ | yes^36^ |
| Protozoa | *Cryptosporidium* | *Cryptosporidium parvum* | If detected, call as *Cryptosporidium parvum* positive | ^37^ | yes |
|  | *Entamoeba histolytica* | *Entamoeba histolytica* | If detected, call as *Entamoeba histolytica* positive | ^38^ | yes |
|  | *Giardia* | *Giardia duodenalis* | If detected, call as *Giardia duodenalis* positive | ^38^ | yes |
| Helminths | *Trichuris* | *Trichuris trichiura* | If detected, call as *Trichuris trichiura* positive | ^39^ | yes |
|  | *Ascaris* | *Ascaris lumbricoides* | If detected, call as *Ascaris lumbricoides* positive | ^40^ | yes |
| Controls | --- | Internal Positive Control | If detected in all rows, assume no inhibition | ^10^ | --- |
|  | --- | MS2 Phage (extraction control) | If detected in all rows, consider extraction successful | ^41^ | --- |

**Quantitative molecular assays: ddPCR.**

For density estimation, we conducted absolute quantification of 12 enteric pathogen targets in high-volume aerosol samples via Droplet Digital PCR (ddPCR; QX200 Droplet Digital PCR System, Bio-Rad, Hercules, CA, USA). Targets included nucleic acids associated with selected viruses (adenovirus A-F, pan-enterovirus, norovirus GI, and norovirus GII), bacteria (*Campylobacter jejuni*, *Shigella*/EIEC, ST-ETEC, and two targets for *Salmonella* spp.), and protozoa (*Cryptosporidium* spp*.* and *Giardia duodenalis*) and are detailed in SI Table 2. We screened all primer, probe, and control sequences in NCBI BLASTn to confirm reported specificities and ordered all control genetic materials from Integrated DNA Technologies (Coralville, IA).

We confirmed assay performance in our specific sample matrix (bioaerosols) by experimentally determining 95% limits of detection (LODs) for each assay using a ten-replicate serial dilution series of positive control material and a probit analysis outlined by Stokdyk et. al.^42,43^ We incubated control materials at 50°C for 20 min, hydrated according to manufacturer instructions, and performed a serial dilution. We vortexed for 15 seconds between every transfer and made sure to pipetting depth was consistent across the dilution. We used these replicates to calculate the 95% LOD which represents the concentration for which the probability of a single ddPCR reaction being positive is 95%. Positive control sequences, primers, probes, and experimentally determined 95% LODs are detailed for each assay in Table S2.

Before ddPCR for RNA targets (detailed in SI Table 2), we performed reverse transcription (RT) of RNA to cDNA with a High Capacity cDNA Reverse Transcription Kit with RNase Inhibitor (ThermoFisher Scientific, Waltham, MA). Following the manufacturer’s instructions, we added 10 μL of nucleic acid extract to 10 μL of RT master mix and loaded the 20 μL reaction on to the thermal cycler with the following conditions: 1) 10 minutes at 25°C, 2) 120 minutes at 37°C, 3) 4 min at 85°C and 5) infinite hold at 4°C. We stored the resulting cDNA at -80°C until further molecular analysis. We assumed 100% efficiency and reverse ddPCR results were then multiplied by a 0.5 factor to account for the 1:2 dilution of extract. In interpreting assays that underwent reverse transcription, we first assessed the positive detection and amplification of the process control spike of RNA virus BRSV via the Inforce3 bovine vaccine in each sample. We stored cDNA at -20°C until analysis within a week.

During ddPCR, the PCR reaction is partitioned into thousands of individual reaction partitions before amplification using QX200 Droplet Generator, sealed using the PX1 PCR Plate Sealer, amplified using the C1000 Touch Thermal Cycler and analyzed at end-point with the QX200 Droplet Reader to enable absolute quantification of target DNA or cDNA (Bio-Rad, Hercules, CA, USA). Unlike qPCR, no standard curve is necessary as targets are quantitatively estimated using a most-probable number technique based on the Poisson distribution and the observed proportion of droplets positive for the target of interest^44^. We conduct manual thresholding based on classification of positive or negative droplets using QuantaSoft (V1.7.4; BioRad, Hercules, CA).

For probe-based assays, we set reaction mixes to a total volume of 20 µL including 0.5 µL each of forward and reverse primer for a final concentration of 900 nM; 0.05 µL of probe for a final concentration of 250 nM; 10 µL of 2X Supermix for Probes (Bio-Rad, Hercules, CA, USA), 5 µL of molecular grade water, and 4 µL of extract. The only assay not using a probe-based assay was *E. coli* (*ybbW*) for which we used EvaGreen chemistry in a total reaction volume of 20 uL that included 1 µL each of forward and reverse primers for final concentrations of 250 nM; 10 µL of 2X EvaGreen Supermix (Bio-Rad, Hercules, CA, USA); 4 µL of molecular grade water, and 4 µL of extract (Supporting Information). We performed each ddPCR experiment using the Bio-Rad QX200 Droplet Digital PCR System and C1000 Touch Thermal Cycler (Bio-Rad, Hercules, CA, USA). On each ddPCR plate, we included two positive controls, two sample blank controls (extracted elution buffer) and two no-template controls using molecular-grade water to control for contamination via human or other error and to detect false positives*.*

**Table S2.** ddPCR assay specs, thermocycling conditions, and 95% limits of detection (95% LOD).

| **Target** | **Reference** | | **Gene** | | **RT** | | **Primers** | | **Probe** | | **Sequence position** | **Amplicon length** | **Thermal Cycling Conditions** | | **95% LOD (gc/reaction)** | | **Gen bank accession** | |
| --- | --- | --- | --- | --- | --- | --- | --- | --- | --- | --- | --- | --- | --- | --- | --- | --- | --- | --- |
| Bovine respiratory syncytial virus | | ^45^ | | *beta-actin* | | Yes | | F:GCAATGCTGCAGGACTAGGTATAAT R:ACACTGTAATTGATGACCCCATTCT | | 5'- /56-FAM/ACCAAGACT/ZEN/TGTATGATGCTGCCAAAGCA/3IABkFQ/ -3' | F: 992-1016  R: 1115-1091  P: 1043-1061 | 124 | | 95 C 10 min; 95 C 30s, 56.6 C 2 min (40x); 98 C 10 min; 4 C hold | | --- | | AF295544.1 |
| *Shigella*/Enteroinvasive *E. coli* (EIEC) | | ^46^ | | *ipaH* | | No | | F:ACCATGCTCGCAGAGAAACT R:TACGCTTCAGTACAGCATGC | | 5'- /5HEX/TGGCGTGTC/ZEN/GGGAGTGACAGC/3IABkFQ/ -3' | F: 1345-1364  R: 1525-1506  P: 1401-1421 | 181 | | 95 C 10 min; 95 C 30s, 58.7 C 2 min (40x); 98 C 10 min; 4 C hold | | 2.02 | | M76445.1 |
| Heat Stabile Enterotoxigenic *E. coli* (ST-ETEC) | | ^47^ | | *STh* | | No | | F:TCCTGAAAGCATGAATAGTAGCAATTAC R:TTAATAGCACCCGGTACAAGCA | | 5-' /56-FAM/ACAACACAATTCACAGCA/3MGBEC/ -3' | F: 171-198  R: 243-222  P: 268-291 | 73 | | 95 C 10 min; 94 C 30s, 54.6 C 1 min (40x); 98 C 10 min; 4 C hold | | 5.07 | | M29255.1 |
| *Campylobacter jejuni* | | ^48^ | | *mapA* | | No | | F: CTGGTGGTTTTGAAGCA  AAGATT R:CAATACCAGTGTCTAAA  GTGCGTTTAT | | 5'- /6FAM/TTGAATTCCAACATCGCTAATGTATAAAAGCCCTTT/MGBNFQ/ -3' | F: 307-329  R: 402-376  P: 330-365 | 95 | | 95 C 10 min; 95 C 30s, 58.7 C 1 min (40x); 98 C 10 min; 4 C hold | | 3.40 | | X80135.1 |
| *Salmonella* sp. | | ^49^ | | *ttr* | | No | | F: CTCACCAGGAGATTAC  AACATGG R: AGCTCAGACCAAAAGT  GACCATC | | 5'- /56-FAM/AAAGTCGGT/ZEN/CTCGCCGTCGGTG/3IABkFQ/ -3' | F: 4287-4309  R: 4381-4359  P: 4336-4357 | 95 | | 95C 10 min; 95 C 30s, 59.9 C 1 min (40x); 98 C 10 min; 4 C hold | | 3.77 | | AF282268 |
| *Salmonella* sp. | | ^50^ | | *invA* | | No | | F: TCGTCATTCCATTACCT  ACC R: AAACGTTGAAAAACTG  AGGA | | 5'- /5HEX/TCTGGTTGA/ZEN/TTTCCTGATCGCA/3IABkFQ/ -3' | F: 167-186  R: 285-234  P: 189-210 | 118 | | 95 C 10 min; 94 C 30 s, 51.2 C 1 min (40x); 98 C 10 min; 4 C hold | | 1.71 | | M90846 |
| *E. coli* | | ^51^ | | *ybbW* | | No | | F: TGATTGGCAAAATCTGG  CCG R:GAAATCGCCCAAATCGCCAT | | n/a | F: 538033-538052  R: 538224-538243 | 211 | | 95 C 10 min; 95 C 30s, 59 C 1.5 min (40x); 4 C 5 min; 90 C 5 min; 4 C hold | | 7.58 | | NC_000913.3 |
| *Giardia duodenalis* | | ^52^ | | *Beta-giardin* | | No | | F:GGCCCTCAAGAGCCTGAAC R:GGGCGATCGTCTCCTTCTC | | 5'- /56-FAM/CTCGAGACAGGCATC/3MGBEC/ -3' | F: 402-420  R: 544-526  P: 268-291 | 143 | | 95 C 10 min; 95 C 30s, 58.7 C 1 min (40x); 98 C 10 min; 4 C hold | | 2.85 | | AY072727 |
| *Cryptosporidum* sp. | | ^4,53^ | | *18S rRNA* | | No | | F:GGGTTGTATTTATTAGATAAAGAACCA R:AGGCCAATACCCTACCGTCT | | 5'- /5HEX/TGACATATCATTCAAGTTTCTGAC/3MGBEC/ -3' | F: 197-223  R: 322-303  P: 268-291 | 80 | | 95 C 10 min; 94 C 30s, 54.6 C 1 min (40x); 98 C 10 min; 4 C hold | | 1.71 | | AF093491.1 |
| norovirus GII | | ^34^ | | *ORF 1-2 junction* | | Yes | | F:CARGARBCNATGTTYAGRTGGATGAG R:TCGACGCCATCTTCATTCACA | | 5'- /56-FAM/TGGGAGGGC/ZEN/GATCGCAATCT/3IABkFQ/ -3' | F: 5003-5028  R: 5100-5080  P: 5048-5067 | 98 | | 95 C 10 min; 95 C 30s, 54.6 C 1 min (40x); 98 C 10 min; 4 C hold | | 4.66 | | AF145896.1 |
| pan-enterovirus | | ^54,55^ | | *EntV F' UTR* | | Yes | | F: CCTCCGGCCCCTGAATG R:ACCGGATGGCCAATCCAA | | 5'- /56-FAM/CGGAACCGA/ZEN/CTACTTTGGGTGTCCGT/3IABkFQ/ -3' | F: 424-440  R: 619-602  P: 512-537 | 196 | | 95 C 10 min; 95 C 30s, 56.2 C 2 min (40x); 98 C 10 min; 4 C hold | | 1.72 | | MW030642.1 |
| norovirus GI | | ^33^ | | *ORF1-ORF2* | | Yes | | F:GCCATGTTCCGNTGGATG R:TCCTTAGACGCCATCATCAT | | /5HEX/TGTGGACAG/ZEN/GAGATCGCAATCTC/3IABkFQ/ | F: 5266-5283  R: 5361-5342  P: 5303-5325 | 96 | | 95 C 10 min; 95 C 30s; 54.6 C 1 min (40x); 98 C 10 min; 4 C hold | | 2.30 | | MG049693.1 |
| adenovirus A-F | | ^27^ | | *hexon*/*JTVX* | | Yes | | F:GGACGCCTCGGAGTACC  TGAG R:ACNGTGGGGTTTCTGAA  CTTGTT | | 5'- /56-FAM/CTGGTGCAG/ZEN/TTCGCCCGTGCCA/3IABkFQ/ -3' | F: 18895-18915  R: 18990-18968  P: 18923: 18944 | 96 | | 95 C 10 min; 95 C 30s, 58.7 C 2 min (40x); 98 C 10 min; 4 C hold | | 4.31 | | AC_000008 |
| MS2 | | ^41^ | | *MS2g1* | | No | | F: TGGCACTACCCCTCTCC  GTATTCACG R:GTACGGGCGACCCCACG  ATGAC | | 5'- /5HEX/CACATCGAT/ZEN/AGATCAAGGTGCCTACAAGC/3IABkFQ/ '3' | F: 160-185  R: 358-237  P: 201-229 | 99 | | 95C 10 min; 95 C 30s, 59.9 C 1 min (40x); 98 C 10 min; 4 C hold | | 3.15 | | NC_001417 |

**Table S3:** Model results for samples in La Paz and Kanpur. In in the linear regression analyses of meteorological variable effects on target density, multivariable models were conducted first to narrow down meaningful variables. Subsequently, individual linear regression models were conducted and only the variables that were the most meaningful are reported.

|  | Relevant Effect Models | | | | | | | | | | | |
| --- | --- | --- | --- | --- | --- | --- | --- | --- | --- | --- | --- | --- |
|  | La Paz | | | | | | Kanpur | | | | | |
|  | Unpaired Two-Samples Wilcoxon Test: Season (rainy or dry) | | Unpaired Two-Samples Wilcoxon Test: Time of day (morning or afternoon) | | Linear regression analysis: effect of temperature in the morning | | Unpaired Two-Samples Wilcoxon Test: Season (rainy or dry) | | Unpaired Two-Samples Wilcoxon Test: Time of day (morning or afternoon) | | Linear regression analysis: effect of relative humidity | |
| Target | p-value | effect size | p-value | effect size | p-value | adjusted R^2^ | p-value | effect size | p-value | effect size | p-value | adjusted R^2^ |
| adenovirus A-F | 0.64 | 0.058 | 0.40 | 0.11 | 0.0043 | 0.18 | 0.16 | 0.20 | 0.81 | 0.034 | 0.39 | -0.0061 |
| ST-ETEC | 0.49 | 0.082 | 0.22 | 0.15 | 0.0050 | 0.18 | 0.090 | 0.23 | 0.49 | 0.097 | 0.68 | -0.021 |
| *Salmonella sp.* | 0.60 | 0.063 | 0.88 | 0.020 | 0.51 | -0.015 | 0.22 | 0.172 | 0.81 | 0.038 | 0.49 | -0.013 |
| *Campylobacter jejuni* | 0.49 | 0.082 | 0.88 | 0.020 | 0.51 | -0.015 | 0.90 | 0.020 | 0.74 | 0.048 | 0.99 | -0.026 |
| pan-enterovirus | 0.39 | 0.10 | 0.13 | 0.19 | 0.34 | -0.0020 | 0.42 | 0.11 | 0.54 | 0.087 | 0.45 | -0.011 |
| EIEC/*Shigella sp.* | 0.13 | 0.18 | 0.87 | 0.022 | 0.023 | 0.11 | 0.27 | 0.16 | 0.42 | 0.12 | --- | --- |
| norovirus GI | 0.39 | 0.10 | 0.75 | 0.041 | 0.028 | 0.10 | 0.000030 | 0.58 | 0.030 | 0.296 | 0.040 | 0.084 |
| norovirus GII | 0.39 | 0.10 | 0.74 | 0.043 | 0.0050 | 0.18 | 0.050 | 0.28 | 0.14 | 0.204 | 0.030 | 0.099 |
| *E. coli* | 0.047 | 0.23 | 0.79 | 0.034 | 0.85 | -0.026 | 0.070 | 0.25 | 0.47 | 0.10 | 0.4 | -0.0067 |

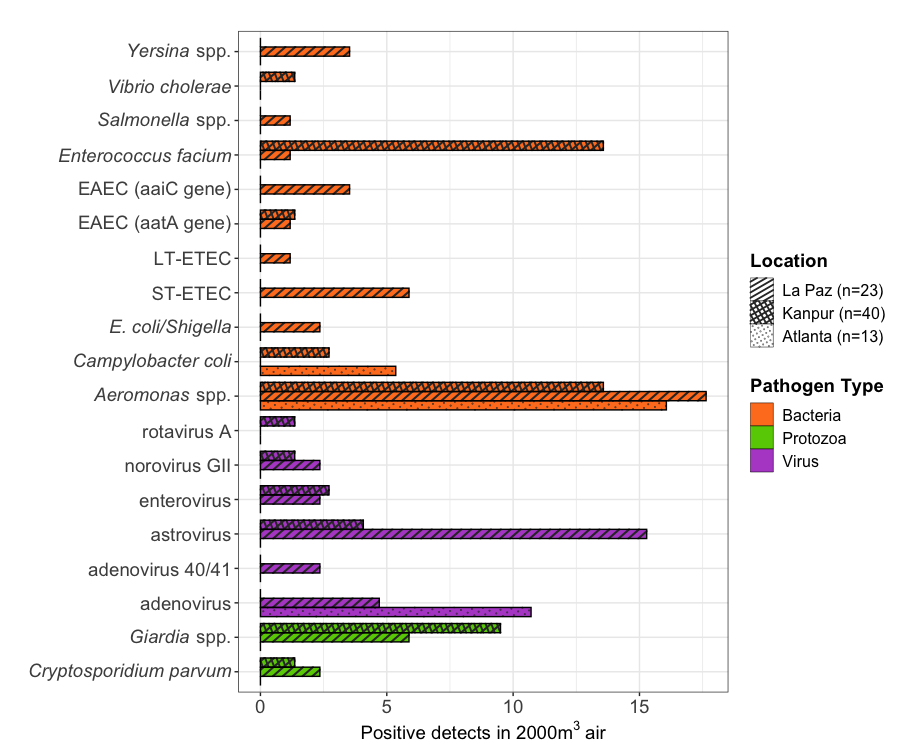

**Figure S1**. TAC detection of enteric pathogen targets in 2000 m^3^_air_, stratified by location and pathogen group.

**Molecular detection of *E. coli* (qPCR and ddPCR), Bolivia.** We detected an average of 2.0×10^4^ (± 3.3×10^4^) gc per m^3^_air_ (n=75) via ddPCR. Through qPCR, we detected any *E. coli* species in 31% of samples (n=26). Of the matched qPCR and ddPCR samples (n=26), we detected *E. coli* through qPCR in 23% of samples that had detectable *E. coli* through ddPCR (Figure S2). We detected culturable *E. coli* in 23% of the samples with detectable *E. coli* when we had data from both methods.

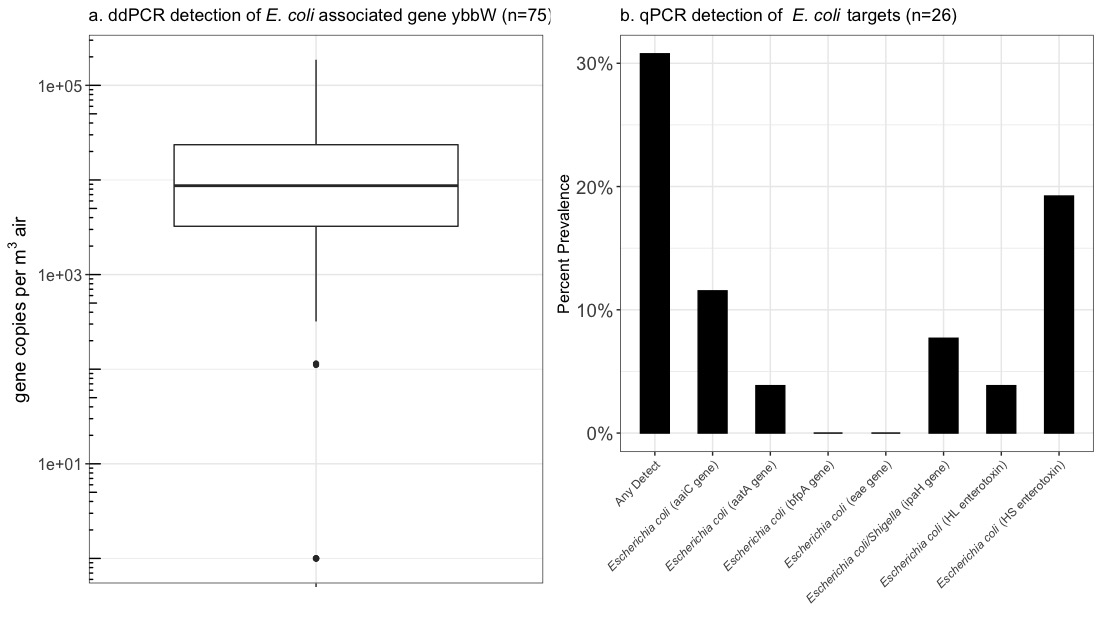

**Figure S2**. (a) We detected through ddPCR the E. coli associated gene ybbW in La Paz. The target was present in 92% of samples. (b) We detected through qPCR multiple E. coli strains and multiple gene targets.

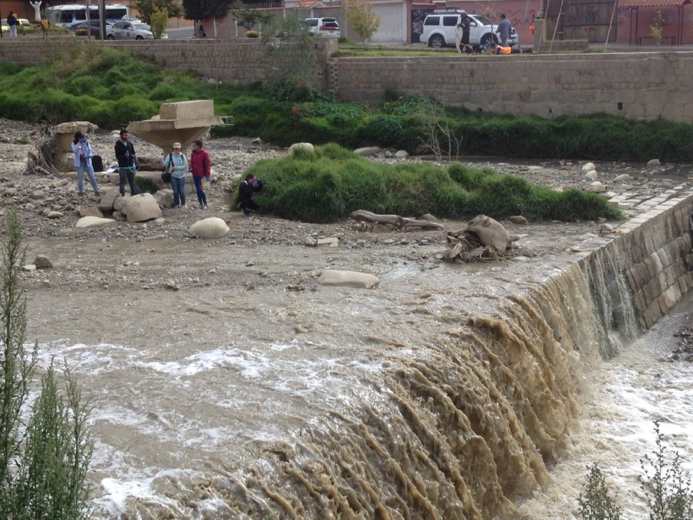

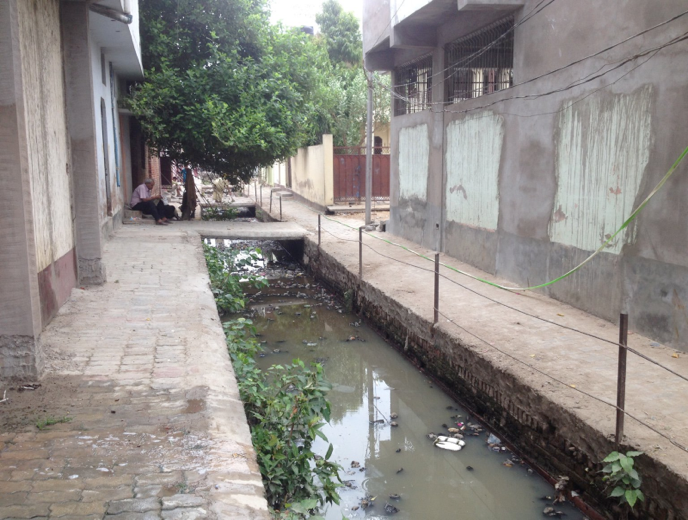

**Figure S3**. Typical open wastewater canals in Kanpur, India (left) and La Paz, Bolivia (right). Photos by Joe Brown.

**Open waste canal water samples.**

To confirm fecal contamination in environmental water sources adjacent to bioaerosol sampling sites in Kanpur, India, we collected 45 mL liquid grab samples from nearby surface waters at 11 OWCs. We analyzed OWC samples via the Luminex xTAG Gastrointestinal-Pathogen Panel (GPP) on Luminex MagPix device (Thermo Scientific™, USA). This multiplex, RT-PCR based assay simultaneously detects the presence/absence of the following microbes that represent important enteric pathogens globally and indicate potential fecal contamination^52–54^: adenovirus 40/41, rotavirus A, norovirus GI/GII, *Salmonella* spp.(including serovars Typhi and Paratyphi), *Campylobacter* spp. (*C. jejuni, C. coli, C. lari*), *Shigella* spp. (S.*boydii*, *S. sonnei, S. flexneri, S. dysenteriae*), *Clostridium difficile* Toxin A/B, enterotoxigenic *Escherichia coli* (ETEC) LT/ST, *E. coli* O157, Shiga-like toxin-producing *E. coli* (STEC) stx1/stx2, *Yersinia enterocolitica, Vibrio cholerae, Giardia duodenalis*, *Entamoeba histolytica*, and *Cryptosporidium* spp. (*C.parvum, C. hominis*). In La Paz, we collected 4 100 mL grab samples from 3 sampling sites on the Choqueyapu river. We analyzed these samples via TAC qPCR (Table S4).

**Table S4**: Presence in OWCs of enteric pathogens tested via the Luminex xTAG GPP multiplex assay.

| ***Kanpur*** | | | ***La Paz*** | | |
| --- | --- | --- | --- | --- | --- |
| ***Number of Samples*** | ***Positive Detects*** | ***Method*** | ***Number of Samples*** | ***Positive detects*** | ***Method*** |
| *11* | *Giardia spp.* (9), ETEC (5), *Campylobacter spp.* (4), *Salmonella* *spp*. (1), Norovirus (1) | GPP | 4 | Pan-adenovirus (2), adenovirus 40/41 (1), pan-astrovirus (3), pan-enterovirus (1), norovirus GI (2), norovirus GII (1), rotavirus A (1), sapovirus I/II/IV (2), sapovirus V (1), *Aeromonas spp.* (4), EAEC (3), EPEC (1), ST-ETEC (3), LT-ETEC (2), EIEC/*Shigella spp.* (2), *E. faecium* (1), STEC (2), *Yersinia spp.* (2), *Giardia duodenalis* (4) | TAC |

**REFERENCES: Supporting Information**

(16)	Ryan, K. J.; Ray, G. C. *Sherris Medical Microbiology: An Introduction to Infectious Diseases*, 4th ed.; McGraw-Hill Medical, 2003.
